# Educational attainment and genetic liability to overweight: Body mass index across the adult life course in females and males

**DOI:** 10.64898/2026.03.07.26347869

**Authors:** María Fernanda Vinueza-Veloz, Ben M. Brumpton, Neil M. Davies, Øyvind E. Næss

## Abstract

**Background and Aim:** Socioeconomically disadvantaged people are more likely to have high body mass index (BMI). However whether socioeconomic position moderates genetic susceptibility to high BMI, and whether this effect differs by sex, remains unclear. We aimed to investigate whether educational attainment (EA) moderates the association between genetic liability for high BMI and BMI trajectories across adulthood in females and males.

**Methods:** We analyzed data from 69,314 participants in the Trøndelag Health Study (HUNT), a population-based cohort with genotyping and repeated BMI measures. A polygenic index for BMI (BMI PGI) was calculated, and participants were categorized by EA level. Using linear mixed-effects models stratified by sex, we tested the interaction between BMI PGI, EA, and age on BMI trajectories.

**Results:** The relationship between BMI PGI and BMI was non-linear, showing a steeper slope in the upper deciles, and was modified by sex (*p*<0.001). Sex-stratified analysis showed that EA moderated the effect of BMI PGI on BMI in females (*p*=0.003) but not in males (*p*=0.089). Among highly educated females, the BMI difference between the top and bottom BMI PGI deciles was-0.99 kg/m² [95%CI: -1.67 to -0.30] smaller than among those with low education. In males, the corresponding difference was -0.16 kg/m² [-0.71 to 0.39]. Genetic influences on BMI trajectories showed consistent age-dependent patterns across all educational groups, though trajectories differed by sex. Females experienced a steady increase in BMI until age 60, after which it declined. Males had an early rapid increase, then stabilization, followed by a slight late-life decline.

**Conclusion:** Higher EA consistently moderates the effect of genetic liability for high BMI in females throughout adulthood, but this protective effect is absent in males. This sex difference suggests that gender-related socioeconomic factors may modulate the expression of BMI-related genetic variants, warranting further investigation into the underlying mechanisms.

## Introduction

Having an unhealthy high body mass index (BMI) represents a major global health challenge that contributes to the burden of chronic diseases and adversely impacts overall well-being and mental health (1,2). Across populations, research consistently reveals a dynamic socioeconomic gradient in the prevalence of high BMI that varies according to a population’s stage in the so-called obesity epidemic (3). During the early stages (Stage 1 and 2), high BMI is more prevalent among socioeconomically advantaged groups, particularly females. However, as the epidemic progresses to Stage 3, this pattern reverses, so that high BMI becomes less common among individuals with a higher socioeconomic position (3).

Among more recent cohorts, this inverse gradient is clearly reflected when socioeconomic position is measured by educational attainment (EA), with higher EA associated with lower BMI, a disparity that likely emerges early in the life course (4). Specifically, children of parents with high EA enter adolescence on lower BMI trajectories, which persist into adulthood (5–7). Yet it is unlikely that these social gradients unfold in isolation from genetic influences, given that genetic factors explain a substantial proportion of BMI variation throughout the life course (8,9). Moreover, the expression of the genetic liability to high BMI is itself likely contingent on sociodemographic factors, including EA (10,11).

Consistent with this, studies have shown that higher EA moderates the association between BMI polygenic index (BMI PGI) and measured BMI, with some evidence suggesting sex differences (12–15). Yet critical questions remain regarding whether this moderating effect shapes BMI trajectories throughout adulthood and, if so, whether these trajectories differ between sexes, as findings on these longitudinal patterns are limited and mixed (15–17). Elucidating sex-specific BMI trajectories is essential because males and females experience distinct biological transitions and gender pressures.

For females, life course events such as pregnancy and menopause are associated with significant physiological changes that directly influence BMI variation (18,19). These biological cycles further intersect with structural constraints, particularly gendered labor markets and caregiving burdens, which tend to generate obesogenic pressures for women (20–22). For males, biological transitions are primarily driven by a gradual decline in androgen levels and a concomitant increase in visceral adiposity (23). These changes occur within a broader context of occupational exposures and culturally prescribed masculine norms, which together shape a distinct, men-specific trajectory toward high BMI (24).

Therefore, to address these questions, we adopt a life-course perspective to test whether the hypothesized moderation effect of EA on genetic liability remains stable, weakens, or strengthens over time as individuals navigate distinct biological and social pathways. Using longitudinal data from the Trøndelag Health Study (HUNT), we examine whether EA moderates the association between genetic liability for high BMI and BMI trajectories across adulthood, and whether these moderating effects differ by sex (25). By integrating genomic data with repeated BMI measurements in this large, population-based cohort, our analysis aims to illuminate how social stratification and sex-specific life-course processes jointly shape the expression of genetic risk for high BMI.

## Methods

### Sample

The HUNT study is a series of population-based health surveys of adults residing in Trøndelag County, Norway. Four surveys have been conducted from 1984 to 2019: HUNT1 (1984–1986), HUNT2 (1995–1997), HUNT3 (2006–2008), and HUNT4 (2017–2019). Participation rates ranged from 89.4% in HUNT1 (77,202 participants) to 54.0% in HUNT4 (56,042 participants). In total, 125,000 individuals have participated in at least one survey wave, and 18,895 individuals participated in all four surveys, providing longitudinal data spanning more than three decades.

### Phenotype data

Information on education was collected in HUNT1, HUNT2, and HUNT4 using the question, “What is your highest level of education?”. Responses were harmonized across surveys and converted into years of schooling (Supplementary Table 1). For each participant, the highest reported number of years across surveys was assigned. Education was then categorized into three groups reflecting the Norwegian system: Low (≤ 10 years, compulsory schooling); middle (11-12 years, upper secondary school); and high (>12 years, tertiary education).

BMI was calculated as weight in kilograms (kg) divided by height in meters (m) squared (kg/m^2^). Measurements were taken with participants wearing light clothes and no shoes. In HUNT1 and HUNT2, height was measured to the nearest centimeter (cm) and weight to the nearest half kg. In HUNT3 and HUNT4, both height and weight were measured to one decimal place. Age was calculated as the time interval between January 1 of year of birth and the date of participation.

### Genotyping

Genotyping was available for 70,517 participants from HUNT2 or HUNT3 (25). Standard HumanCoreExome arrays (HumanCoreExome12 v1.0 and v1.1) were used to genotype 12 864 samples, and a customised HumanCoreExome array (UM HUNT Biobank v1.0) was used for the remainder. Quality control was performed separately for each array. The overall call rate exceeded 99%. Imputation was performed on samples of recent European ancestry using Minimac3 (v2.0.1) from a merged reference panel constructed from the Haplotype Reference Consortium panel (release version 1.1) and a merged local reference panel based on 2201 whole-genome sequenced HUNT participants (26–28).

### Polygenic index

We constructed a BMI PGI based on the PGI derived and validated by Khera et al. (29). The original Khera et al. PGI comprised approximately 2 million genetic variants, excluding ambiguous strand variants. For the present analysis, we refined this set by excluding variants with a minor allele frequency (MAF) < 0.05 and R^2^ < 0.8 in the HUNT sample. The resulting index consisted of 1,837,194 variants. Individual BMI PGI were then calculated using PLINK 2.0 by multiplying each variant’s allelic dosage by its estimated coefficient and summing across all variants (30). For analysis, we ranked the BMI PGI into deciles.

### Ethical approval

The present study was approved by the Mid-Norway Regional Committee for Medical and Health Research Ethics (REK Midt 2015/1197). All participants provided written informed consent, including consent for genetic analyses.

### Statistical analysis

We conducted all analyses using R statistical software (31). A series of linear mixed-effects models with random intercepts and random slopes was fitted to account for the repeated BMI measurements (Models 1, 2, and 3). The models were specified with the lmer() function from the lmerTest package. All estimates were adjusted for the first 20 genetic principal components and for genotyping batch. Age was standardised to z-scores (mean = 0, SD = 1) before inclusion in the models.

To accommodate flexible, non-linear relationships among the BMI PGI, age, and phenotypic BMI, we modelled both the BMI PGI and age as cubic B-splines using the bs() function from the splines package. The splines were given four degrees of freedom; for cubic splines this corresponds to three interior knots placed at the 25ᵗʰ, 50ᵗʰ, and 75ᵗʰ percentiles of the variable’s distribution, with boundary knots at the observed minimum and maximum values. Model 1 included sex as an effect modifier, while Models 2 and 3 were fitted separately for females and males.

- Model 1 estimated the association between BMI PGI and BMI, including EA as covariate and the following interaction terms: BMI PGI × age, BMI PGI × sex, BMI PGI × HUNT-survey, EA × sex, HUNT-survey × sex, and age × sex.
- Model 2 tested whether EA modifies the effect of BMI PGI on BMI by adding a BMI PGI × EA interaction.
- Model 3 extended this by testing a three-way interaction, adding a BMI PGI × EA × age term.

To evaluate the statistical significance of the interaction terms in all three models, we performed Type III F-tests with Satterthwaite’s approximation for the degrees of freedom, as implemented in the anova() function from lmerTest. This approach tests whether all coefficients that compose a given interaction term (e.g., the 16 coefficients for the BMI PGI × age spline interaction) are jointly different from zero, thereby providing a single test of effect modification.

We generated predicted values with the emmeans package for each combination of categorical variables, holding continuous variables at their mean values; the resulting predictions were then averaged to obtain the reported estimates. Using Model 2, we calculated the average predicted BMI for three informative BMI PGI deciles (1st, 5th, and 10th) across levels of education. Within each education level, we compared average predicted BMI between PGI deciles (1 vs 5, 1 vs 10, and 5 vs 10) by computing the differences and testing these contrasts. To control for multiple testing across decile contrasts and education levels, *p*-values were adjusted with Tukey’s method. An analogous procedure was applied to the results from Model 3.

## Results

The sample comprised 69,314 individuals who participated in at least one HUNT survey. Among these, 59,615 (86.0%) participated in at least two HUNT surveys and 36,481 (52.6%) in at least three. In all surveys, the proportion of female participants was higher than that of male participants. Mean age increased sequentially across surveys, although the variance in age was lowest in the most recent survey. The average BMI and standard deviation (SD) increased across surveys from 24.90 kg/m^2^ (SD, 3.70) to 27.5 kg/m^2^ (SD, 4.50) (Table 1). The proportion of participants achieving a high level of education, equivalent to having attended college or university, increased from 11.20% to 37.00% across surveys. The BMI PGI explained 6.54% of the variance in measured BMI.

**Table 1.** Characteristics of the study population. The sample included 69 314 individuals who participated in at least one HUNT survey. Educations was not measured in HUNT3. *Abbreviations and symbology: n, number; %, percentage; SD, standard deviation; kg, kilograms; m, meters; BMI, body mass index; Low, ≤ 10 years of education; Middle, >10 and ≤ 12 years of education; High, >12 years of education*.

|  | **HUNT1**<br>**(n = 43 845)** | **HUNT2**<br>**(n = 56 538)** | **HUNT3**<br>**(n = 48 746)** | **HUNT4**<br>**(n = 36 764)** |
| --- | --- | --- | --- | --- |
| **Sex** |  |  |  |  |
| Female, n (%) | 23 095 (53) | 30 017 (53) | 26 525 (54) | 20 332 (55) |
| Male, n (%) | 20 750 (47) | 26 521 (47) | 22 221 (46) | 16 432 (45) |
| **Age (years)** |  |  |  |  |
| Mean (SD) | 44.30 (14.0) | 50.20 (16.8) | 53.80 (16.0) | 62.60 (13.8) |
| Missing, n | 0 | 26 | 0 | 0 |
| **BMI (kg/m<sup>2</sup>)** |  |  |  |  |
| Mean (SD) | 24.90 (3.7) | 26.30 (4.1) | 27.20 (4.4) | 27.5 (4.5) |
| Missing, n | 634 | 594 | 254 | 1,035 |
| **Education** |  |  |  |  |
| Low, n (%) | 20 455 (57.0) | 19 229 (36.0) |  | 5100 (14.0) |
| Middle, n (%) | 11 685 (32.0) | 23 561 (44.0) |  | 17 651 (49.0) |
| High, n (%) | 4 061 (11.2) | 10 897 (19.8) |  | 13 195 (37.0) |
| Missing, n | 7644 | 2851 |  | 818 |

The relationship between BMI PGI and observed BMI was non-linear, showing a steeper slope in the upper deciles. Sex modified this association (F(4, 63 241) = 20.76, *p* < 0.001). In females, BMI increased progressively but non-uniformly across PGI deciles, from 24.67 kg/m² [95% CI: 24.55 to 29.80] in the first decile to 29.27 kg/m² [29.14 to 29.40] in the tenth decile, a total increase of 4.60 kg/m² [4.28 to 4.91]. In males, a similar pattern was observed, though the curve was somewhat flatter, particularly in the middle deciles. Male BMI rose from 25.40 kg/m² [25.26 to 25.53] to 29.10 kg/m² [28.96 to 29.23], corresponding to an increase of of 3.70 kg/m² [3.37 to 4.03]. The increase in females was 0.90 kg/m² [0.55 to 1.24] higher than in males (*p* < 0.001) (Supplementary Figure 1).

### Educational attainment as an effect modifier of the genetic liability to high BMI

We found that EA modifies the association between BMI PGI and BMI in females (F(8, 32,823) = 2.89, *p* = 0.003) but not in males (F(8, 30,181) = 1.72, *p* = 0.089) (Figure 1). In females, the BMI difference between the top and bottom BMI PGI deciles (1 vs. 10) was larger among those with low (4.98 kg/m² [5.51 to 5.45]) in comparison to those with middle (4.70 kg/m² [4.34 to 5.07]), or high (3.99 kg/m² [3.55 to 3.44]) EA. In males, the absolute BMI differences across BMI PGI deciles were consistently smaller and with little evidence of an educational gradient (low: 3.66 kg/m² [3.25 to 4.08]); middle: (3.88 kg/m² [3.61 to 4.16]); high: (3.50 kg/m² [3.14 to 3.87]) (Supplementary Figure 2). The BMI difference between the top and bottom PGI deciles was -0.99 kg/m² [-1.67 to -0.30] lower among females with a high EA compared to those with a low EA (*p* = 0.002). In contrast, among males, the corresponding difference was -0.16 kg/m² [-0.71 to 0.39] (*p* = 0.774) (Figure 2).

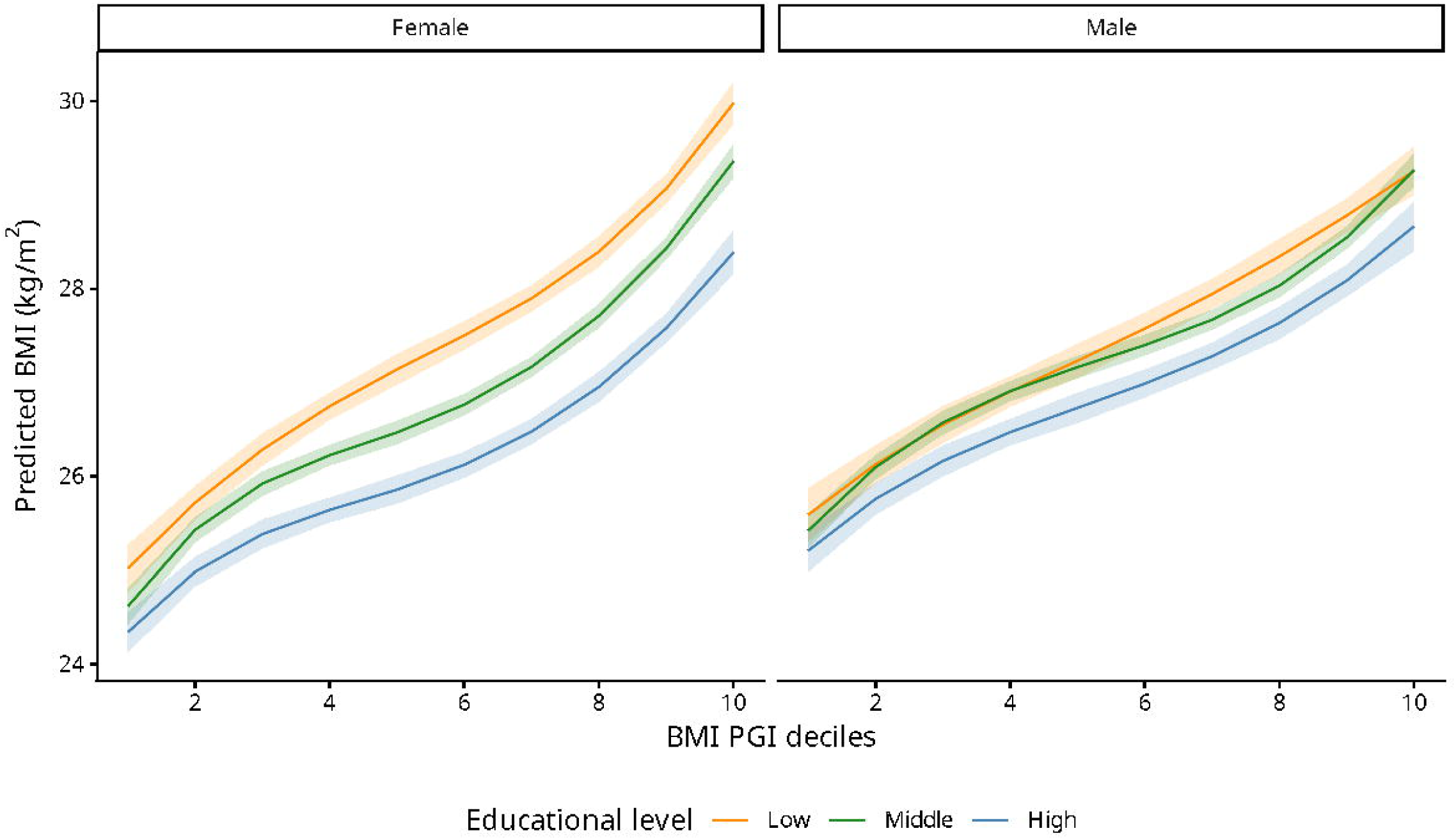

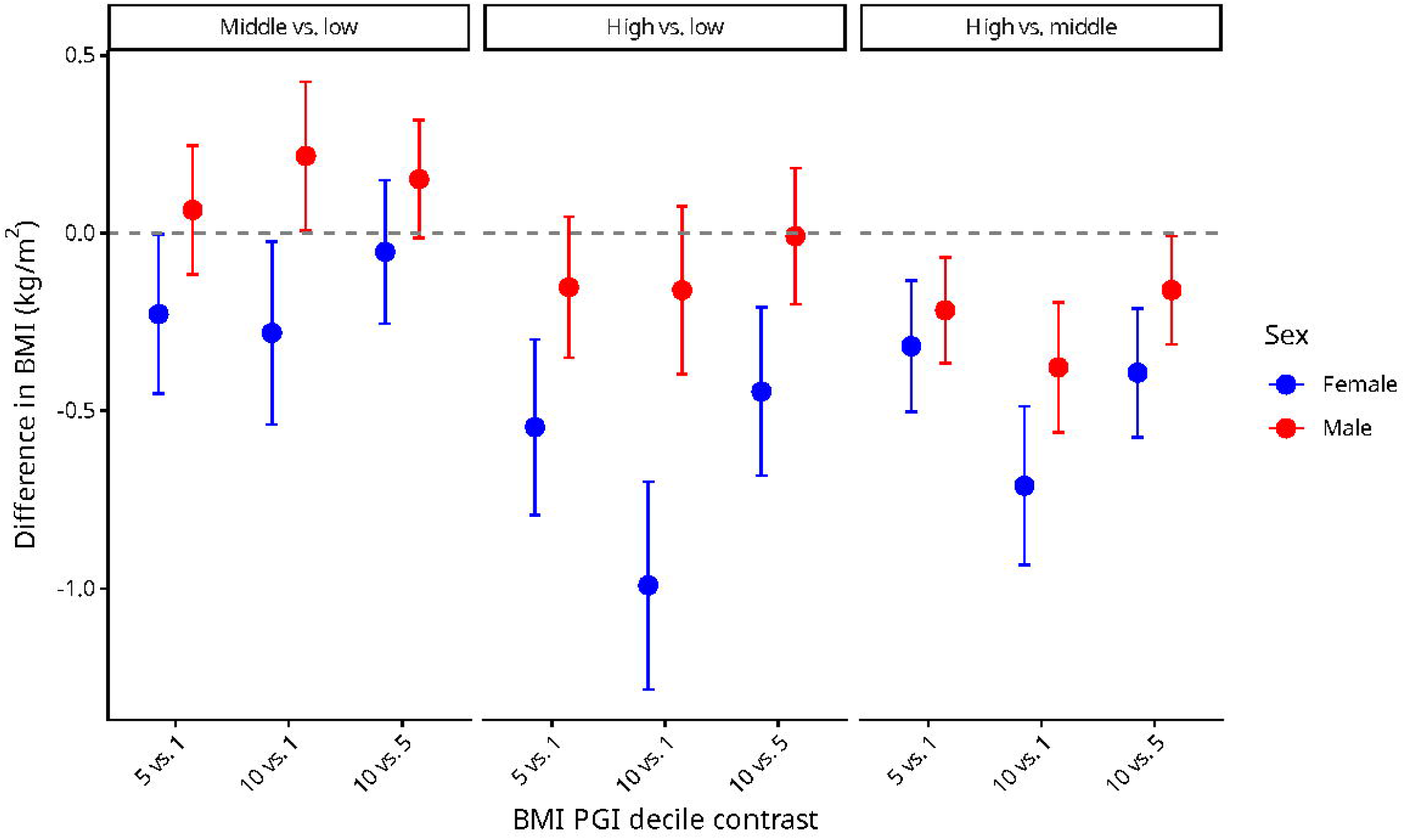

### Educational attainment as an effect modifier of the genetic liability to high BMI across adulthood

To assess whether EA modifies the genetic influence on BMI trajectories across adulthood, we tested a three-way interaction between BMI PGI, age, and EA. There was little evidence of a three-way interaction in females (F(32, 53970) = 1.34, *p* = 0.094) and males (F(32, 46359) = 1.35, *p* = 0.091). Nevertheless, we found a significant two-way interaction between genetic liability and age in both sexes (females: F(16, 77219) = 3.12, *p* < 0.001); males: (F(16, 63174) = 4.80, *p* < 0.001). Together, these results suggest that the association between PGI and BMI varies across adulthood and is consistent across all educational groups for both females and males.

In females, BMI steadily increased from age 20 to 60 and then declined, with the magnitude of change varying by genetic liability, a pattern that held consistently across all educational levels (Figure 3). Among females in the highest BMI PGI decile, BMI at age 60 was 3.58 kg/m² [2.59 to 4.56] higher than at age 20 for those with high education, and 4.07 kg/m² [2.31 to 5.83] for those with low education. There was no difference between high- and low-educated females over this 40-year period (−0.49 kg/m² [-2.02 to 1.04]; *p* = 0.732). From age 60 to 80, both groups experienced BMI declines of -1.23 kg/m² [-2.14 to -0.31] and -1.38 kg/m² [-1.85 to -0.91]. Again, no difference was observed between the two educational levels (0.15 kg/m² [-0.58 to 0.88]; *p* = 0.876). A similar, although less pronounced, pattern was observed for females in the lowest BMI PGI deciles (Supplementary Table 2 and Supplementary Figure 3).

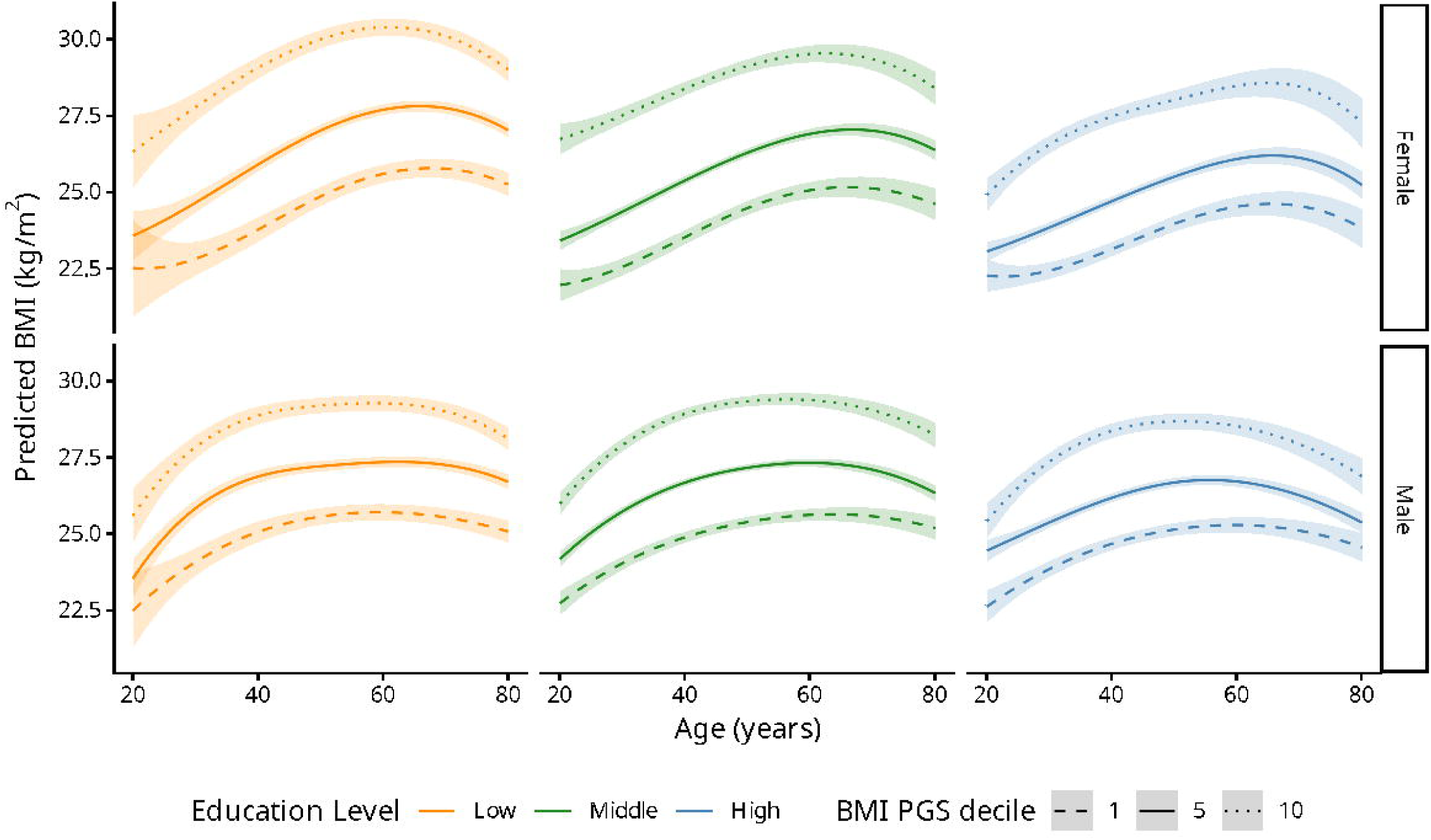

In males, BMI increased most rapidly during early adulthood (ages 20-40), remained stable until around age 60, and then declined modestly, a pattern that was consistent across all educational levels (Figure 3). Males in the highest BMI PGI decile showed greater gains between ages 20 and 30 than between ages 30 and 40 (Supplementary Table 2). Between ages 20 and 30, highly educated males gained 1.96 kg/m² [1.21 to 2.70], compared with 2.24 kg/m² [1.13 to 3.36] among low-educated males; however, there were few differences between educational groups (−0.29 kg/m² [-1.35 to 0.78], *p* = 0.803). Between ages 60 and 80, both educational groups experienced declines in BMI (highly educated males: -1.64 kg/m² [-2.32 to -0.97]; low-educated males: -1.16 kg/m² [-1.59 to -0.73]). Again, no difference was observed between the two educational levels (−0.49 kg/m² [-1.06 to 0.08]; *p* = 0.113). A similar, though less pronounced pattern, was observed for males in the lowest BMI PGI deciles (Supplementary Table 2 and Supplementary Figure 4).

## Discussion

Our longitudinal analysis demonstrates that higher EA moderates the expression of genetic liability for high BMI across adulthood, but this protective effect is evident only in females. Using repeated measures from the HUNT study, we found that the association between the BMI PGI and phenotypic BMI was weaker among highly educated females than among those with lower education, a buffering effect that remained stable from young adulthood to older age. In contrast, we observed no moderating effect by EA in males. Beyond the moderating effect of EA, we also observed that genetic liability influenced BMI trajectories in both sexes, though these followed distinct life-course patterns. In females, BMI increased steadily until late midlife, then declined. In males, BMI rose rapidly in early adulthood, then stabilized before a slight decline in later life.

### Previous studies

Studies investigating whether genetic liability for high BMI interacts with social factors have yielded mixed results, likely reflecting differences in methodological approaches, population contexts, and sample sizes. For instance, Tyrrell et al. (2017) found that socioeconomic deprivation (as measured by the Townsend index) amplifies genetic effects on BMI in the most deprived individuals (12). Tommerup et al. (2021) also found evidence that socioeconomic position moderates genetic liability for high BMI, but the patterns were complex, varying by sex, age group, and which socioeconomic measure was used (15). In contrast, Bann et al. (2022) found little evidence of interaction, instead showing additive effects, as both genetic liability for high BMI and socioeconomic position appear to independently influence BMI (16).

Two studies have specifically examined whether EA moderates genetic liability for high BMI (13,14). Barcellos et al. (2018) used a natural experiment—the 1972 raising of the school-leaving age in the United Kingdom—to investigate whether additional schooling reduces genetic disparities in body size (13). Based on data from approximately 250 000 UK Biobank participants, they constructed a body size index combining BMI, body fat percentage, and waist-hip ratio. They reported that an additional year of schooling reduced the gap in unhealthy body size between those in the top and bottom terciles of genetic liability from 20 to 6 percentage points, with larger improvements observed among individuals at higher genetic liability. Complementing these findings, Komulainen et al. (2018) used prospective data from 2441 participants in the Cardiovascular Risk in Young Finns Study followed over 31 years to examine whether EA moderates genetic effects on BMI trajectories from ages 18 to 49 (14). At age 49, the difference in average BMI between low and high genetic liability groups was 3.3 kg/m² for those with low education and 2.4 kg/m² for those with high education.

### Sex differences

Whether the moderating effect of EA on genetic liability for higher BMI differs by sex remains largely unexplored. Most previous studies have treated sex as a covariate, implicitly assuming that any moderating effect of EA operates similarly in females and males. An exception is the Finnish cohort examined by Silventoinen et al. (2024), who tested the moderating effect of three indicators of socioeconomic position, including EA (17). Their study revealed two key patterns: i) females experienced steeper socioeconomic gradients in BMI, yet ii) the moderating influence of EA on genetic liability was comparable for both sexes. A complementary approach was taken by Calvin et al. (2019) in their analysis of UK Biobank participants, who stratified their models by sex (32). Aligning with our findings, their sex-stratified analyses revealed that college education moderated genetic effects on BMI in females but not in males.

The observed sex difference in how EA buffers genetic liability for higher BMI warrants consideration of underlying mechanisms. Several pathways could explain why EA moderates the effects of BMI PGI in females but not in males. One plausible hypothesis is that the sex difference reflects sex-specific differential responses to the same socioeconomic environment. This formulation would suggest that females and males, when exposed to the same socioeconomic conditions, respond differently, perhaps due to inherent biological differences, distinct psychological dispositions, or divergent health behaviors. This interpretation, however, overlooks a more fundamental asymmetry; the educational gap itself is smaller in men (17,33). Across the adult lifespan, women with low education have approximately 2–3 BMI units higher than their college-educated peers; among men, the corresponding gap is only about 1 BMI unit.

Because men’s BMI varies little across educational groups, the incremental resource gain from additional schooling offers modest protective potential. For women, higher education fundamentally shifts exposure to less obesogenic contexts, thereby providing a larger buffer against genetic liability for high BMI. Moreover, although education is often assumed to confer identical resources to all genders, this is unlikely. A university degree confers income, occupational prestige, and health knowledge to all people irrespective of gender. But these formal equivalents operate within profoundly different life circumstances. For example, a highly educated woman must still navigate a world shaped by gender inequality, caregiving expectations, and reproductive imperatives.

### Strengths and limitations

Strengths of our study include its large sample size, long follow-up, and repeated BMI measurements, which enabled us to model life-course trajectories in both females and males. This longitudinal design was essential for testing the gene-environment interaction between genetic liability and EA across adulthood. Nevertheless, some limitations should be acknowledged. First, the cohort was observed during a period of substantial population-wide increase in BMI. This poses a challenge for life-course analyses because age and period effects are difficult to disentangle. Although we observed age-dependent patterns these trajectories could partly reflect secular trends rather than ageing effects. Previous research in the HUNT population showing increasing BMI PGI effect sizes from the 1960s to the 2000s across all age groups supports this concern (11). Second, BMI does not distinguish between fat and lean mass, which follow different trajectories across the life course—lean mass typically declines in older ages (34). If the BMI PGI has larger effects on fat than lean mass, its influence may be more pronounced in older age groups. Although more precise measures such as dual-energy X-ray absorptiometry (DXA) were not available in HUNT, previous research suggests that complex adiposity measures do not substantially alter associations with major health outcomes compared to BMI (35).

Third, as with any long-term prospective study, missing data and loss to follow-up—particularly in the more recent HUNT waves—may introduce bias. However, a key strength of our analytical approach is the use of linear mixed-effects models, which are robust to missing data under the missing at random (MAR) assumption. These models include all available data points from participants regardless of whether they provided complete data across all waves, thereby minimizing bias and maximizing statistical power compared to complete-case analyses. Fourth, socioeconomic position is multidimensional, and results may differ if other dimensions beyond EA —such as household income or occupational status—are considered, particularly if these capture different stages of the life course. However, our focus on EA was both deliberate and well-justified. EA is a relatively stable measure that also serves as a key determinant of later-life opportunities, making it a particularly meaningful indicator of the social environment across the life course. Finally, our analyses are based on individuals of European ancestry, and findings may not apply to other populations.

### Conclusions

This longitudinal analysis of the HUNT Study cohort demonstrates that higher educational attainment moderates the expression of genetic liability for high BMI across adulthood, but this protective effect is evident only in females. Among women, the association between genetic predisposition and BMI was weaker in those with higher education, and this buffering effect remained stable from young adulthood through older age. In males, despite clear genetic influences on BMI trajectories, we found little evidence of moderation by education. These findings contribute to a growing understanding of how socioeconomic factors and sex differences jointly shape the expression of genetic risk. They also highlight the need to investigate the specific social and biological mechanisms underlying these sex-specific patterns and their implications for obesity prevention strategies.

## Acknowledgments

The Trøndelag Health Study (HUNT) is a collaboration between HUNT Research Center (Faculty of Medicine and Health Sciences, NTNU, Norwegian University of Science and Technology), Trøndelag County Council, Central Norway Regional Health Authority, and the Norwegian Institute of Public Health. The genotyping in HUNT was financed by the National Institutes of Health; University of Michigan; the Research Council of Norway; the Liaison Committee for Education, Research and Innovation in Central Norway; and the Joint Research Committee between St Olavs hospital and the Faculty of Medicine and Health Sciences, NTNU. We thank Paul R. Jones for his valuable contributions to the analysis and manuscript preparation.

## Funding/Support

MFVV and ØEN are funded by The Research Council of Norway, Project Inequalities in non-communicable diseases: Indirect selection or social causation (INDI-INEQ #287347). BMB is funded by the Research Council of Norway (FRIMEDBIO #287347) and by grants from the Liaison Committee for Education, Research and Innovation in Central Norway and the Joint Research Committee between St Olavs Hospital and the Faculty of Medicine and Health Sciences, NTNU. NMD is supported by The Research Council of Norway (#295989) and National Institute of Mental Health (MH130448).

## Role of the Funder

The funders had no role in the design, analysis, interpretation of the data, drafting of the manuscript, review, or final approval of the version to be published, nor in the decision to submit the manuscript for publication.

## Ethical standards

The authors assert that all procedures contributing to this work comply with the ethical standards of the relevant national and institutional committees on human experimentation and with the Helsinki Declaration of 1975, as revised in 2008.”

## Information on author access to data

MFVV and BMB had full access to all of the data in the study and take responsibility for the integrity of the data and the accuracy of the data analysis.

## Authors’ contributions

Conception and design: MFVV and ØEN. Analysis: MFVV. Interpretation of data for the manuscript: MFVV, BMB, NMD and ØEN. Drafting of the manuscript: MFVV and ØEN. Critical review of the manuscript for important intellectual content: MFVV, BMB, NMD and ØEN. Final approval of the version to be published: MFVV, BMB, NMD and ØEN. Agreement to be accountable for all aspects of the work: MFVV, BMB, NMD and ØEN.

## Data availability

Researchers associated with Norwegian research institutes can apply for the use of HUNT material: data and samples - given approval by a Regional Committee for Medical and Health Research Ethics. Researchers from other countries can also apply in cooperation with a Norwegian Principle Investigator. Information for data access can be found at https://www.ntnu.edu/hunt/data.

## Conflict of interest disclosures

MFVV, BMB, NMD and ØEN declare no conflict of interest to disclose.

## Notes

### Competing Interest Statement

The authors have declared no competing interest.

### Funding Statement

MFVV and OEN are funded by The Research Council of Norway, Project Inequalities in non-communicable diseases: Indirect selection or social causation (INDI-INEQ #287347). BMB is funded by the Research Council of Norway (FRIMEDBIO #287347) and by grants from the Liaison Committee for Education, Research and Innovation in Central Norway and the Joint Research Committee between St Olavs Hospital and the Faculty of Medicine and Health Sciences, NTNU. NMD is supported by The Research Council of Norway (#295989) and National Institute of Mental Health (MH130448).

## References

1. GBD 2015 Obesity Collaborators, Afshin A, Forouzanfar MH, Reitsma MB, Sur P, Estep K, et al. Health Effects of Overweight and Obesity in 195 Countries over 25 Years. N Engl J Med. 2017 Jul 6;377(1):13–27. doi:10.1056/NEJMoa1614362 PubMed PMID: 28604169; PubMed Central PMCID: PMC5477817.

2. Lingvay I, Cohen RV, Roux CW le, Sumithran P. Obesity in adults. The Lancet. 2024 Sep 7;404(10456):972–87. doi:10.1016/S0140-6736(24)01210-8 PubMed PMID: 39159652.

3. Jaacks LM, Vandevijvere S, Pan A, McGowan CJ, Wallace C, Imamura F, et al. The obesity transition: stages of the global epidemic. Lancet Diabetes Endocrinol. 2019 Mar;7(3):231–40. doi:10.1016/S2213-8587(19)30026-9 PubMed PMID: 30704950; PubMed Central PMCID: PMC7360432.

4. Kinge JM, Strand BH, Vollset SE, Skirbekk V. Educational inequalities in obesity and gross domestic product: evidence from 70 countries. J Epidemiol Community Health. 2015 Dec;69(12):1141–6. doi:10.1136/jech-2014-205353 PubMed PMID: 26179449.

5. Muthuri SK, Onywera VO, Tremblay MS, Broyles ST, Chaput JP, Fogelholm M, et al. Relationships between Parental Education and Overweight with Childhood Overweight and Physical Activity in 9–11 Year Old Children: Results from a 12-Country Study. PLoS One. 2016 Aug 24;11(8):e0147746. doi:10.1371/journal.pone.0147746 PubMed PMID: 27557132; PubMed Central PMCID: PMC4996501.

6. Bae D, Wickrama KAS, O’Neal CW. Social consequences of early socioeconomic adversity and youth BMI trajectories: Gender and race/ethnicity differences. Journal of Adolescence. 2014 Aug 1;37(6):883–92. doi:10.1016/j.adolescence.2014.06.002

7. Bann D, Johnson W, Li L, Kuh D, Hardy R. Socioeconomic inequalities in childhood and adolescent body-mass index, weight, and height from 1953 to 2015: an analysis of four longitudinal, observational, British birth cohort studies. Lancet Public Health. 2018 Apr;3(4):e194–203. doi:10.1016/S2468-2667(18)30045-8 PubMed PMID: 29571937; PubMed Central PMCID: PMC5887082.

8. Silventoinen K, Jelenkovic A, Sund R, Hur YM, Yokoyama Y, Honda C, et al. Genetic and environmental effects on body mass index from infancy to the onset of adulthood: an individual-based pooled analysis of 45 twin cohorts participating in the COllaborative project of Development of Anthropometrical measures in Twins (CODATwins) study123. Am J Clin Nutr. 2016 Aug;104(2):371–9. doi:10.3945/ajcn.116.130252 PubMed PMID: 27413137; PubMed Central PMCID: PMC4962159.

9. Silventoinen K, Jelenkovic A, Sund R, Yokoyama Y, Hur YM, Cozen W, et al. Differences in genetic and environmental variation in adult BMI by sex, age, time period, and region: an individual-based pooled analysis of 40 twin cohorts. Am J Clin Nutr. 2017 Aug;106(2):457–66. doi:10.3945/ajcn.117.153643 PubMed PMID: 28679550; PubMed Central PMCID: PMC5525120.

10. Silventoinen K, Jelenkovic A, Latvala A, Yokoyama Y, Sund R, Sugawara M, et al. Parental education and genetics of body mass index from infancy to old age: a pooled analysis of 29 twin cohorts. Obesity (Silver Spring). 2019 May;27(5):5. doi:10.1002/oby.22451 PubMed PMID: 30950584; PubMed Central PMCID: PMC6478550.

11. Brandkvist M, Bjørngaard JH, Ødegård RA, Åsvold BO, Sund ER, Vie GÅ. Quantifying the impact of genes on body mass index during the obesity epidemic: longitudinal findings from the HUNT Study. BMJ. 2019 Jul 3;366:l4067. doi:10.1136/bmj.l4067 PubMed PMID: 31270083; PubMed Central PMCID: PMC6607203.

12. Tyrrell J, Wood AR, Ames RM, Yaghootkar H, Beaumont RN, Jones SE, et al. Gene-obesogenic environment interactions in the UK Biobank study. Int J Epidemiol. 2017 Apr 1;46(2):559–75. doi:10.1093/ije/dyw337 PubMed PMID: 28073954; PubMed Central PMCID: PMC5837271.

13. Barcellos SH, Carvalho LS, Turley P. Education can reduce health differences related to genetic risk of obesity. Proc Natl Acad Sci U S A. 2018 Oct 16;115(42):E9765–72. doi:10.1073/pnas.1802909115 PubMed PMID: 30279179; PubMed Central PMCID: PMC6196527.

14. Komulainen K, Pulkki-Råback L, Jokela M, Lyytikäinen LP, Pitkänen N, Laitinen T, et al. Education as a moderator of genetic risk for higher body mass index: prospective cohort study from childhood to adulthood. Int J Obes (Lond). 2018 Apr;42(4):866–71. doi:10.1038/ijo.2017.174 PubMed PMID: 28757641.

15. Tommerup K, Ajnakina O, Steptoe A. Genetic propensity for obesity, socioeconomic position, and trajectories of body mass index in older adults. Sci Rep. 2021 Oct 13;11(1):20276. doi:10.1038/s41598-021-99332-7 PubMed PMID: 34645866; PubMed Central PMCID: PMC8514538.

16. Bann D, Wright L, Hardy R, Williams DM, Davies NM. Polygenic and socioeconomic risk for high body mass index: 69 years of follow-up across life. PLoS Genet. 2022 Jul;18(7):e1010233. doi:10.1371/journal.pgen.1010233 PubMed PMID: 35834443; PubMed Central PMCID: PMC9282556.

17. Silventoinen K, Lahtinen H, Kilpi F, Morris TT, Davey Smith G, Martikainen P. Socio-economic differences in body mass index: the contribution of genetic factors. Int J Obes. 2024 May;48(5):741–5. doi:10.1038/s41366-024-01459-w

18. Linné Y, Dye L, Barkeling B, Rössner S. Weight development over time in parous women—The SPAWN study—15 years follow-up. Int J Obes. 2003 Dec;27(12):1516–22. doi:10.1038/sj.ijo.0802441

19. Guo SS, Zeller C, Chumlea WC, Siervogel RM. Aging, body composition, and lifestyle: the Fels Longitudinal Study. Am J Clin Nutr. 1999 Sep;70(3):405–11. doi:10.1093/ajcn/70.3.405 PubMed PMID: 10479203.

20. Kim B, Thomsen MR, Nayga RM, Goudie A. The effect of gender-specific labor market conditions on children’s weight. Health Econ Rev. 2021 Dec 2;11:44. doi:10.1186/s13561-021-00345-9 PubMed PMID: 34855042; PubMed Central PMCID: PMC8641227.

21. Stall NM, Shah NR, Bhushan D. Unpaid Family Caregiving—The Next Frontier of Gender Equity in a Postpandemic Future. JAMA Health Forum. 2023 Jun 9;4(6):e231310. doi:10.1001/jamahealthforum.2023.1310

22. Lee S, Colditz GA, Berkman LF, Kawachi I. Caregiving and risk of coronary heart disease in U.S. women: A prospective study. American Journal of Preventive Medicine. 2003 Feb 1;24(2):113–9. doi:10.1016/S0749-3797(02)00582-2

23. Anawalt BD, Matsumoto AM. Aging and androgens: Physiology and clinical implications. Rev Endocr Metab Disord. 2022 Dec 1;23(6):1123–37. doi:10.1007/s11154-022-09765-2

24. Monaghan LF. Body Mass Index, masculinities and moral worth: men’s critical understandings of ‘appropriate’ weight-for-height. Sociology of Health & Illness. 2007;29(4):584–609. doi:10.1111/j.1467-9566.2007.01007.x

25. Brumpton BM, Graham S, Surakka I, Skogholt AH, Løset M, Fritsche LG, et al. The HUNT study: A population-based cohort for genetic research. Cell Genom. 2022 Oct 12;2(10):10. doi:10.1016/j.xgen.2022.100193 PubMed PMID: 36777998; PubMed Central PMCID: PMC9903730.

26. Das S, Forer L, Schönherr S, Sidore C, Locke AE, Kwong A, et al. Next-generation genotype imputation service and methods. Nat Genet. 2016 Oct;48(10):1284–7. doi:10.1038/ng.3656 PubMed PMID: 27571263; PubMed Central PMCID: PMC5157836.

27. McCarthy S, Das S, Kretzschmar W, Delaneau O, Wood AR, Teumer A, et al. A reference panel of 64,976 haplotypes for genotype imputation. Nat Genet. 2016 Oct;48(10):1279–83. doi:10.1038/ng.3643 PubMed PMID: 27548312; PubMed Central PMCID: PMC5388176.

28. Zhou W, Fritsche LG, Das S, Zhang H, Nielsen JB, Holmen OL, et al. Improving power of association tests using multiple sets of imputed genotypes from distributed reference panels. Genet Epidemiol. 2017 Dec;41(8):744–55. doi:10.1002/gepi.22067 PubMed PMID: 28861891; PubMed Central PMCID: PMC6324190.

29. Khera AV, Chaffin M, Wade KH, Zahid S, Brancale J, Xia R, et al. Polygenic Prediction of Weight and Obesity Trajectories from Birth to Adulthood. Cell. 2019 Apr 18;177(3):587–596.e9. doi:10.1016/j.cell.2019.03.028 PubMed PMID: 31002795; PubMed Central PMCID: PMC6661115.

30. Chang CC, Chow CC, Tellier LC, Vattikuti S, Purcell SM, Lee JJ. Second-generation PLINK: rising to the challenge of larger and richer datasets. Gigascience. 2015;4:7. doi:10.1186/s13742-015-0047-8 PubMed PMID: 25722852; PubMed Central PMCID: PMC4342193.

31. R Core Team. R: A Language and Environment for Statistical Computing [Internet]. Vienna: R Foundation for Statistical Computing; 2021. Available from: https://www.r-project.org/

32. Calvin CM, Hagenaars SP, Gallacher J, Harris SE, Davies G, Liewald DC, et al. Sex-specific moderation by lifestyle and psychosocial factors on the genetic contributions to adiposity in 112,151 individuals from UK Biobank. Sci Rep. 2019 Jan 23;9(1):363. doi:10.1038/s41598-018-36629-0

33. Yang YC, Walsh CE, Johnson MP, Belsky DW, Reason M, Curran P, et al. Life-course trajectories of body mass index from adolescence to old age: Racial and educational disparities. Proceedings of the National Academy of Sciences. 2021 Apr 27;118(17):e2020167118. doi:10.1073/pnas.2020167118

34. Bann D, Johnson W, Li L, Kuh D, Hardy R. Socioeconomic Inequalities in Body Mass Index across Adulthood: Coordinated Analyses of Individual Participant Data from Three British Birth Cohort Studies Initiated in 1946, 1958 and 1970. PLoS Med. 2017 Jan;14(1):e1002214. doi:10.1371/journal.pmed.1002214 PubMed PMID: 28072856; PubMed Central PMCID: PMC5224787.

35. Taylor AE, Ebrahim S, Ben-Shlomo Y, Martin RM, Whincup PH, Yarnell JW, et al. Comparison of the associations of body mass index and measures of central adiposity and fat mass with coronary heart disease, diabetes, and all-cause mortality: a study using data from 4 UK cohorts1234. The American Journal of Clinical Nutrition. 2010 Mar 1;91(3):547–56. doi:10.3945/ajcn.2009.28757

36. Goodpaster BH, Park SW, Harris TB, Kritchevsky SB, Nevitt M, Schwartz AV, et al. The loss of skeletal muscle strength, mass, and quality in older adults: the health, aging and body composition study. J Gerontol A Biol Sci Med Sci. 2006 Oct;61(10):1059–64. doi:10.1093/gerona/61.10.1059 PubMed PMID: 17077199.

