## Supplementary figures and images for "Educational attainment and genetic liability to overweight: Body mass index across the adult life course in females and males"

### Supplementary Figure 1

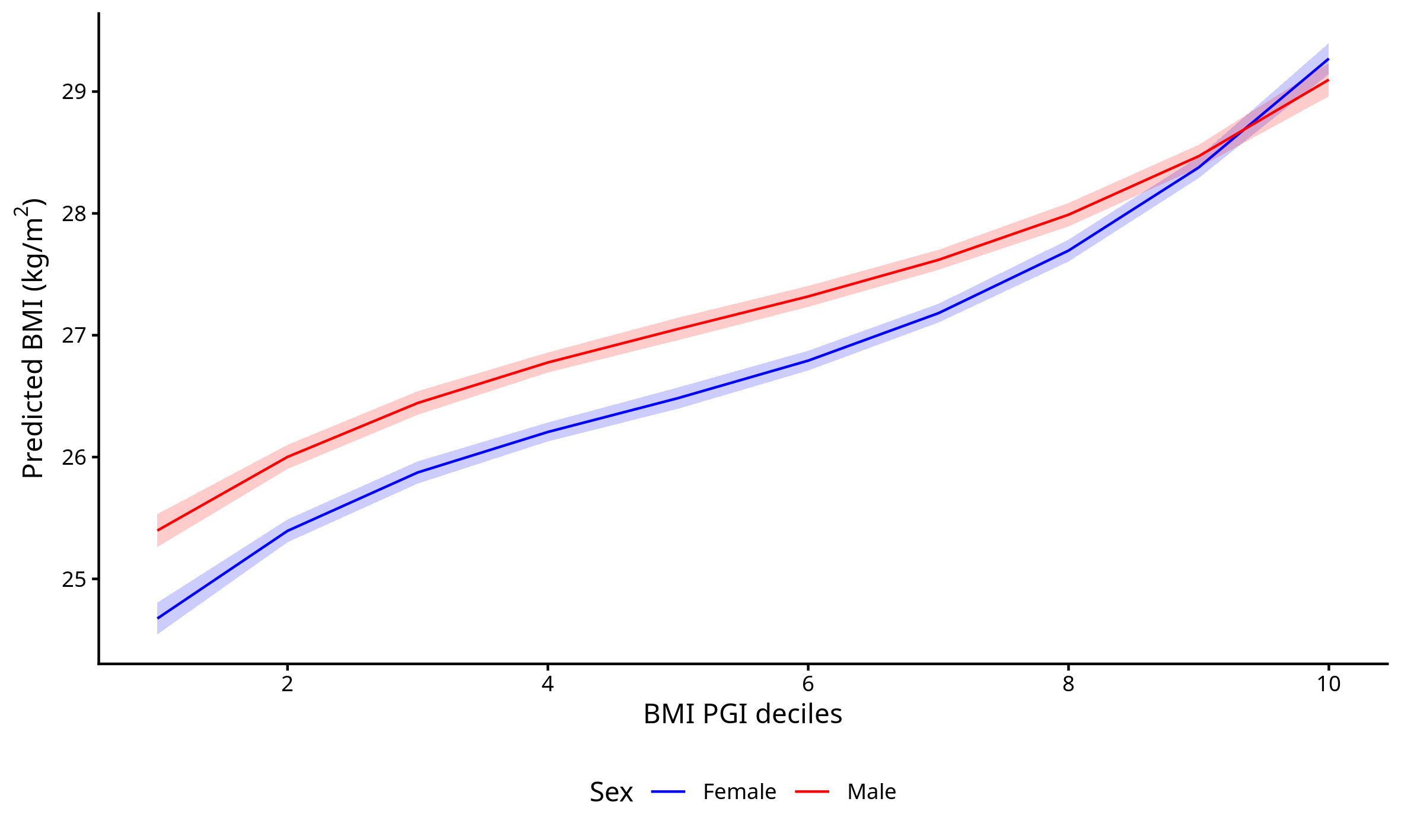

### Supplementary Figure 1

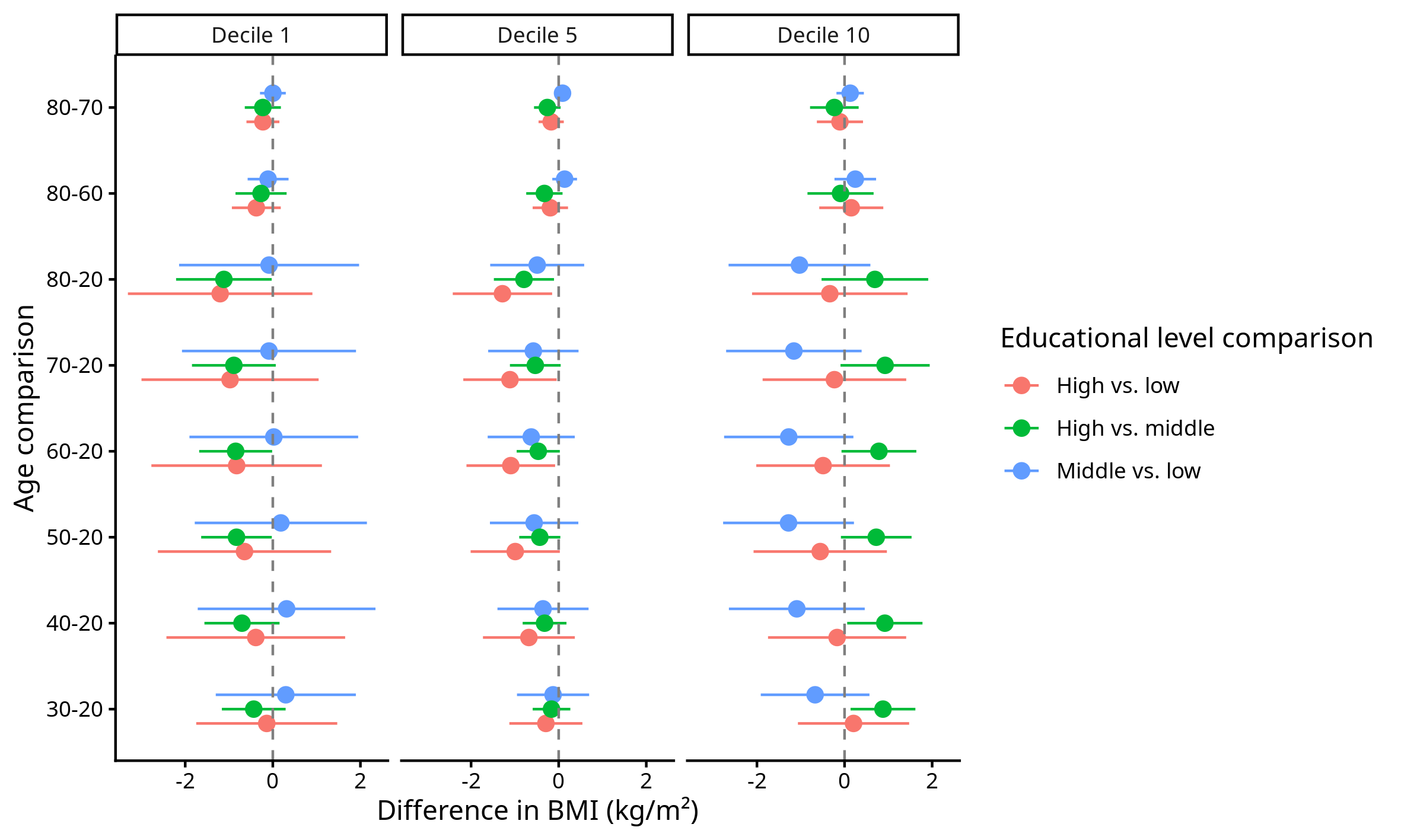

### Supplementary Figure 2

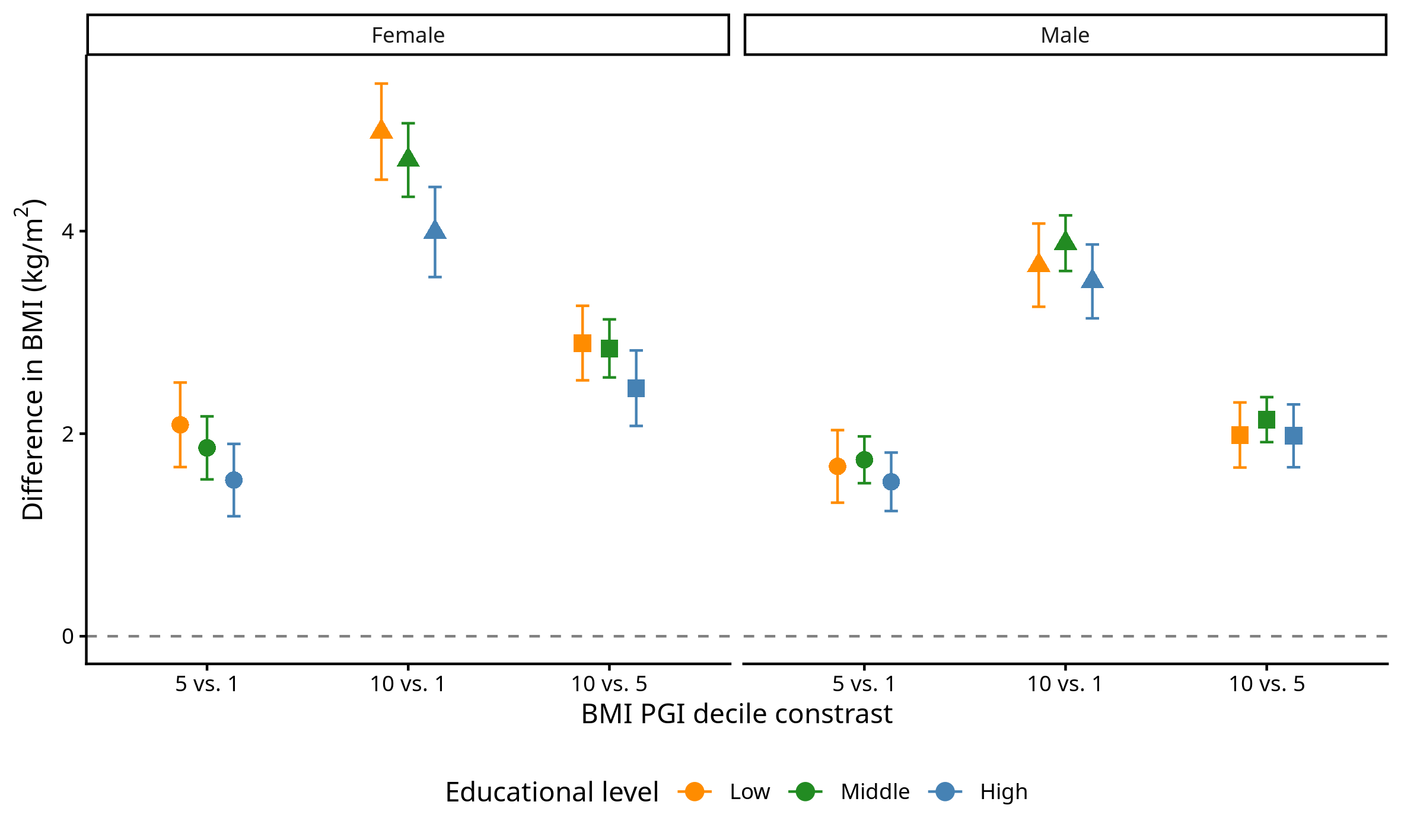

### Supplementary Figure 4

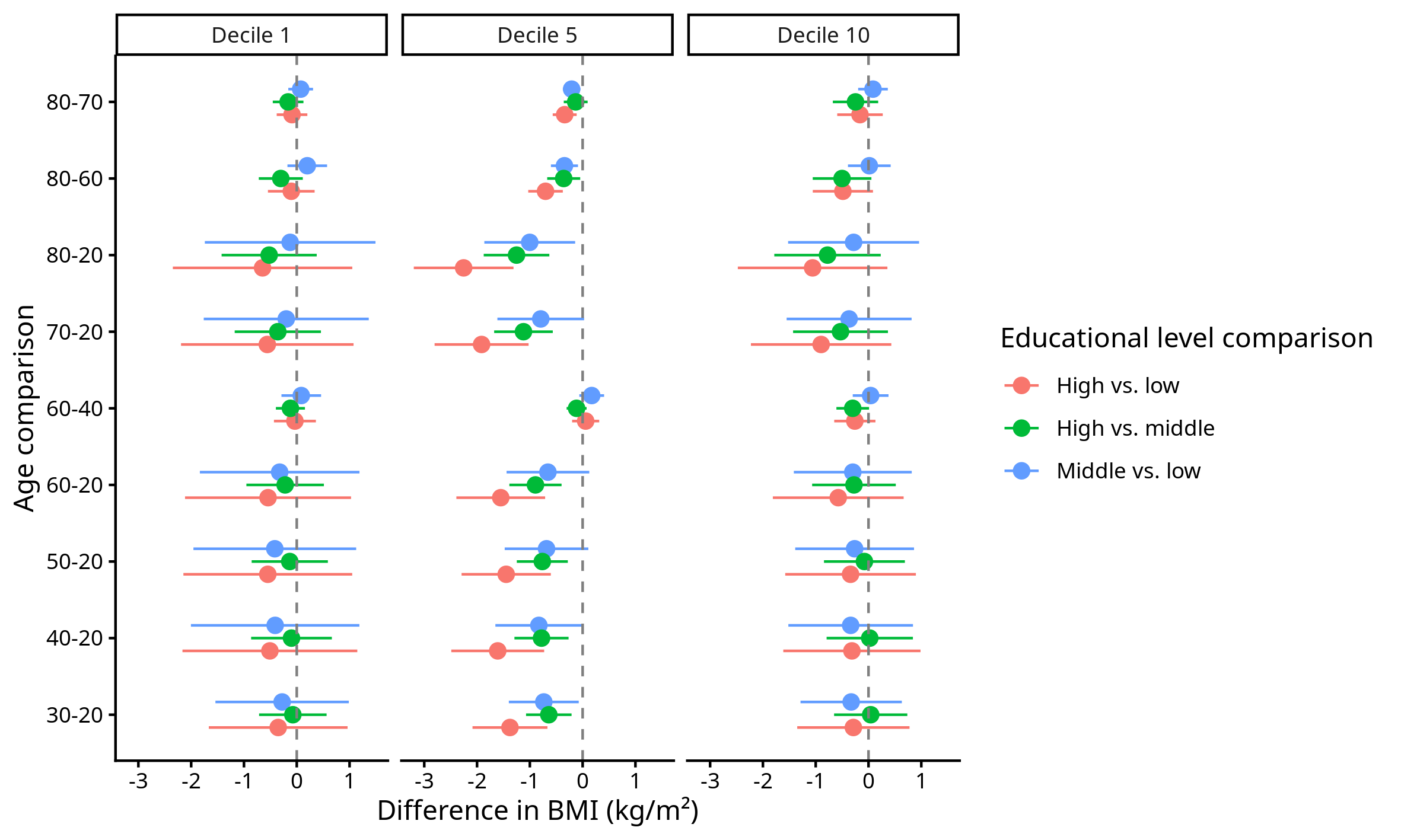
