## Supplementary Table 1 for "Educational attainment and genetic liability to overweight: Body mass index across the adult life course in females and males"

**Supplementary Table 1.** Categories of attained education in the three HUNT surveys.

| Duration of education | HUNT1 | HUNT2 | HUNT4 |
|---|---|---|---|
| 10 Years | 7 years' primary school or less | | |
| | Middle school | | |
| | 9 years' compulsory primary and lower secondary school | | |
| | 10 years' primary and lower secondary school | Primary school 7–10 years, continuation school, folk high school | 9–10 years' compulsory primary and lower secondary school |
| 12 Years | 1 or 2 years at upper secondary school | High school, intermediate school, vocational school, 1–2 years' high school | 1 or 2 years of academic or vocational school |
| | General Certificate of Education, commercial college or sixth form college | | 3 years of academic or vocational school |
| | | University qualifying examination, junior college, A levels | 3–4 years' vocational school/apprentice (upper secondary/sixth form college) |
| 16 Years | College or university, less than 4 years | University or other post-secondary education, less than 4 years | College or university, less than 4 years |
| 17 Years | College or university, 4 years or more | University/college, 4 years or more | College or university, 4 years or more |
